# What innovations help with the recruitment and retention of ambulance staff: a rapid evidence summary

**DOI:** 10.1101/2022.11.29.22282890

**Authors:** Deborah Edwards, Judit Csontos, Elizabeth Gillen, Judith Carrier, Ruth Lewis, Alison Cooper, Adrian Edwards

## Abstract

Ambulance waiting times across the UK have increased in recent years. The numbers of ambulance staff leaving services across the UK is increasing every year. Strategies to help recruit and retain all ambulance staff, including paramedics are important. This rapid evidence summary aimed to investigate what innovations can help with their recruitment and retention.

Eight primary studies were identified:

**Recruitment:** Evidence from a UK survey suggests that factors negatively influencing paramedic recruitment include competitive job market, lack of locally trained professionals, and newly qualified professionals starting with higher debt. Evidence from the US suggests that factors supporting recruitment concern future paramedics wanting to enter a caring profession or an exciting job. Additionally, strategies to recruit emergency medical technicians need to include the motivational aspects of growth, advancement, recognition, and responsibility. Evidence indicates that factors hindering recruitment of emergency medical technicians and/ or paramedics include rural working, and ambulance services not seen as a primary career path.

**Retention:** Evidence from a UK survey suggests that pay, reward, stress and workload are factors that hinder paramedic retention. Evidence recommends retention strategies for paramedics, such as reviewing banding, improving work conditions and career progression, changing the way ambulances are dispatched to calls, and providing retention premiums. Evidence from the US suggests that pay, benefits, opportunities for advancement, continuous professional development, burnout, stress, workload, nearing retirement and career change are factors that influence retention of emergency medical technicians and/ or paramedics. Evidence from Thailand suggests that remuneration and professionalism are factors supporting paramedic retention.

More up-to-date information is needed on the recruitment and retention of ambulance staff in UK settings. Further research providing a more detailed investigation of factors influencing recruitment and retention may be useful. The development or implementation of future strategies to help the recruitment and retention of paramedics and emergency medical technicians should be accompanied by a planned impact evaluation.

**Funding statement:** The Wales Centre For Evidence Based Care was funded for this work by the Wales Covid-19 Evidence Centre, itself funded by Health & Care Research Wales on behalf of Welsh Government.

**Wales COVID-19 Evidence Centre (WCEC) Rapid Evidence Summary:** *What innovations help with the recruitment and retention of ambulance staff: a rapid evidence summary Report number – RES00050 (November 2022):* **Rapid Evidence Summary Details**
Review conducted by
The Wales Centre For Evidence Based Care

Review Team

▪ Deborah Edwards
▪ Judit Csontos
▪ Liz Gillen
▪ Judith Carrier

**Review submitted to the WCEC on:** 24^th^ November 2022
**Stakeholder consultation meeting: 14**^**th**^ **November 2022**
**Rapid Evidence Summary report issued by the WCEC in**: December 2022
**WCEC Team:** Ruth Lewis, Adrian Edwards, Alison Cooper and Micaela Gal involved in drafting the Topline Summary and editing.

This review should be cited as
RES00050, Wales COVID-19 Evidence Centre. A rapid evidence summary of what innovations help with the recruitment and retention of ambulance staff. November 2022

Disclaimer
The views expressed in this publication are those of the authors, not necessarily Health and Care Research Wales. The WCEC and authors of this work declare that they have no conflict of interest.

**What innovations help with the recruitment and retention of ambulance staff: a rapid evidence summary Report number – RES00050 (November 2022):** **TOPLINE SUMMARY**
What is a Rapid Evidence Summary?
Our Rapid Evidence Summaries (RES) are designed to provide an interim evidence briefing to inform further work and provide early access to key findings. They are based on a limited search of key resources and the assessment of abstracts. Priority is given to studies representing robust evidence synthesis. No quality appraisal or evidence synthesis are conducted, and the summary should be interpreted with caution.

Who is this summary for?
Welsh Ambulance Service NHS Trust

Background / Aim of Rapid Evidence Summary
Ambulance waiting times across the UK have increased in recent years, with emergency service performance targets missed. Reasons for the decreasing performance include increasing demand, problems with moving patients through the system, and workforce issues. The numbers of ambulance staff leaving services across the UK is increasing every year with the most acute retention problems affecting paramedics. Strategies to help recruit and retain all ambulance staff, including paramedics are important, therefore this rapid evidence summary aims to investigate what innovations can help with their recruitment and retention.

Key Findings
Eight primary studies were identified.
Extent of the evidence base

▪ Quantitative descriptive surveys (n=6) and qualitative studies (n=2)
▪ Studies were from USA (n=6), **UK (n=1)** and Thailand (n=1).

Recency of the evidence base

▪ The review included evidence available up until October 2022. Included studies were published between 2005 and 2021 with the UK study published in 2015.

Summary of findings

▪ Participants were paramedics (n=3), emergency medical technicians (EMTs) (n=2), paramedics and EMTs (n=2), and emergency medical service directors (n=1).
▪ Studies focused on factors and strategies influencing ambulance staff recruitment (n=5) and/ or retention (n=7).

Recruitment

▪ Evidence from a **UK survey** suggests that factors that negatively influence **paramedic** recruitment include **competitive job market, lack of locally trained professionals, and newly qualified professionals starting with higher debt**.
▪ Evidence from the US suggests that factors supporting recruitment concern **future paramedics** wanting to enter a caring profession or an exciting job. Additionally, strategies to recruit **EMTs** need to include the motivational aspects of growth, advancement, recognition, and responsibility.
▪ Evidence from the US indicates that factors that hinder recruitment of **EMTs and/ or paramedics** include rural working, and ambulance services not seen as a primary career path.
▪ There were **no studies** that evaluated the **effectiveness of innovations or strategies to improve ambulance staff recruitment**.

Retention

▪ Evidence from a **UK survey** suggests that **pay, reward, stress and workload** are factors that hinder **paramedic** retention.
▪ Evidence from a **UK survey** recommends retention strategies for **paramedics**, such as reviewing **banding, improving work conditions and career progression, changing the way ambulances are dispatched to calls, and providing retention premiums**.
▪ Evidence from the US suggests that pay, benefits, opportunities for advancement, continuous professional development, burnout, stress, workload, nearing retirement and career change are factors that influence retention and job satisfaction of **EMTs and/ or paramedics**.
▪ Evidence from Thailand suggests that remuneration and professionalism are factors supporting **paramedic** retention.
▪ There were **no studies** that evaluated the **effectiveness of innovations or strategies to improve ambulance staff retention**.

Implications for practice and research

▪ More up-to-date information is needed on the recruitment and retention of ambulance staff in UK settings.
▪ Further research that provides a more detailed investigation of factors influencing recruitment and retention may be useful, in particular regarding reasons for leaving the profession.
▪ The development or implementation of future strategies to help the recruitment and retention of paramedics and EMTs should be accompanied by a planned impact evaluation.

## 1. CONTEXT / BACKGROUND

Ambulance waiting times across the United Kingdom (UK) have increased in recent years, with emergency service performance targets set by governments missed. Ambulance calls are categorised according to severity of the health problem, leading to three main groups, namely red, amber, and green (Senedd Cymru 2022). Red calls are immediately life-threatening health issues, such as cardiac arrests (Senedd Cymru 2022). Amber calls are described as serious, but not immediately life-threatening conditions, including chest pains, fractures, and stroke (Senedd Cymru 2022). Green refers to non-urgent calls, such as minor injuries and aches, or a person recovering from fainting (Senedd Cymru 2022). Response time should be within eight minutes for red calls, but no similar time limits are set for amber and green. Just 8% of 999 emergency ambulance calls are for people with life-threatening illnesses or injuries, which Eaton et al. (2020) suggests indicates that a large proportion of patients access the ambulance service with lower-acuity presentations. The Welsh Government’s performance target for ambulance services aims to respond to 65% of red calls within the intended eight minutes (Welsh Government 2022b). However, in recent months this performance target has not been met. For example, ambulance services arrived to 50% of red calls within eight minutes in September 2022, one of the lowest percentages on record (Welsh Government 2022b). Reasons for the decreasing performance include increasing demand, problems with moving patients through the system, and workforce issues (Audit Wales 2022).

Paramedics are qualified healthcare professionals, registered with the Health and Care Professions Council (HCPC) they can apply clinical and invasive procedures within a pre-clinical environment, and are initially employed at band 5, progressing to band 6 after two years with opportunities to undertake further training and progress to more senior roles (Welsh Ambulance Service NHS Trust 2022). In the UK, since 2006 the paramedic role has started expanding with new opportunities opening up, such as gaining Bachelor Degree qualification in paramedic practice (Woollard 2006). Standards for paramedic registration and programmes leading to registration are regulated by the HCPC in conjunction with the College of Paramedics, and in 2018 the level of qualification for paramedics’ registration in the UK was raised from Certificate of Higher Education to ‘Bachelor Degree with Honours’ (HCPC 2018). At the point of qualification paramedics are expected to be skilled in several areas including managing exacerbation of long-term conditions, assessing patients presenting with mental ill health and assessing and referring patients with social care needs, in addition to managing a range of undifferentiated urgent care presentations (Eaton et al. 2018, Eaton et al. 2020). Within the UK the NHS Long Term Plan (NHS 2019) opened up opportunities for paramedics to work across a variety of healthcare settings including primary care (Eaton et al. 2020). The systematic review undertaken by Eaton et al. (2020) notes that The College of Paramedics has made distinctions between ‘paramedics’, ‘specialist paramedics’, and ‘advanced paramedics’, and advocated against the use of the term ‘emergency care practitioner’ to describe paramedics in extended roles. Other members of the ambulance team who provide first contact care include and emergency care assistants who work alongside paramedics and are trained to provide pre-clinical care at a band 3 or 4, and emergency medical technicians (EMTs) paid at band 4 who support paramedics but may also operate as a single responder (Welsh Ambulance Service NHS Trust 2022, NHS Health Careers 2022).

Paramedic numbers have shown a steady increase over the years in England (data up to November 2021) when it reached its highest with 17,142 full-time staff members (Nuffield Trust 2022). In Wales, full-time ambulance staff numbers, including emergency medical technicians (EMTs), paramedics, advanced practitioners, and managers, showed a similar trend with an overall increase detected (data up to March 2022) (StatsWales 2022). However, from April 2022 numbers of paramedics in England have started to decline (Nuffield Trust 2022), no recent data is available for Wales. Reports suggest that there is a need for additional ambulance staff in Wales, with the Welsh Government vowing to invest into recruiting 100 more frontline workers (Welsh Government 2022a). The College of Paramedics (2019b) included in its strategic aim “to promote the profession as a career”, although no further information was provided to show how this would be achieved.

It has been reported that the numbers of ambulance staff leaving ambulance services across the UK is increasing every year with the most acute retention problems being paramedics (UNISON, Unite, and GMB 2015). A survey conducted for the College of Paramedics in 2016 (which is only available as an abstract) indicated several reasons why ambulance staff leave the profession (College of Paramedics 2019a). These included a lack of career and development, poor job satisfaction / work experience, management issues and pay and benefits (College of Paramedics 2019a). Furthermore, a recently conducted qualitative evidence synthesis found that the perceived heroism of the paramedic role and the ‘rescue myths’ often influence students’ perception, leading to mis-matches between expectations and reality of the work, affecting retention (Rees et al. 2022). Strategies to help recruit and retain all ambulance staff, including paramedics are important, therefore this rapid evidence summary aims to investigate what innovations can help with their recruitment and retention.

## 2. RESEARCH QUESTION(S)

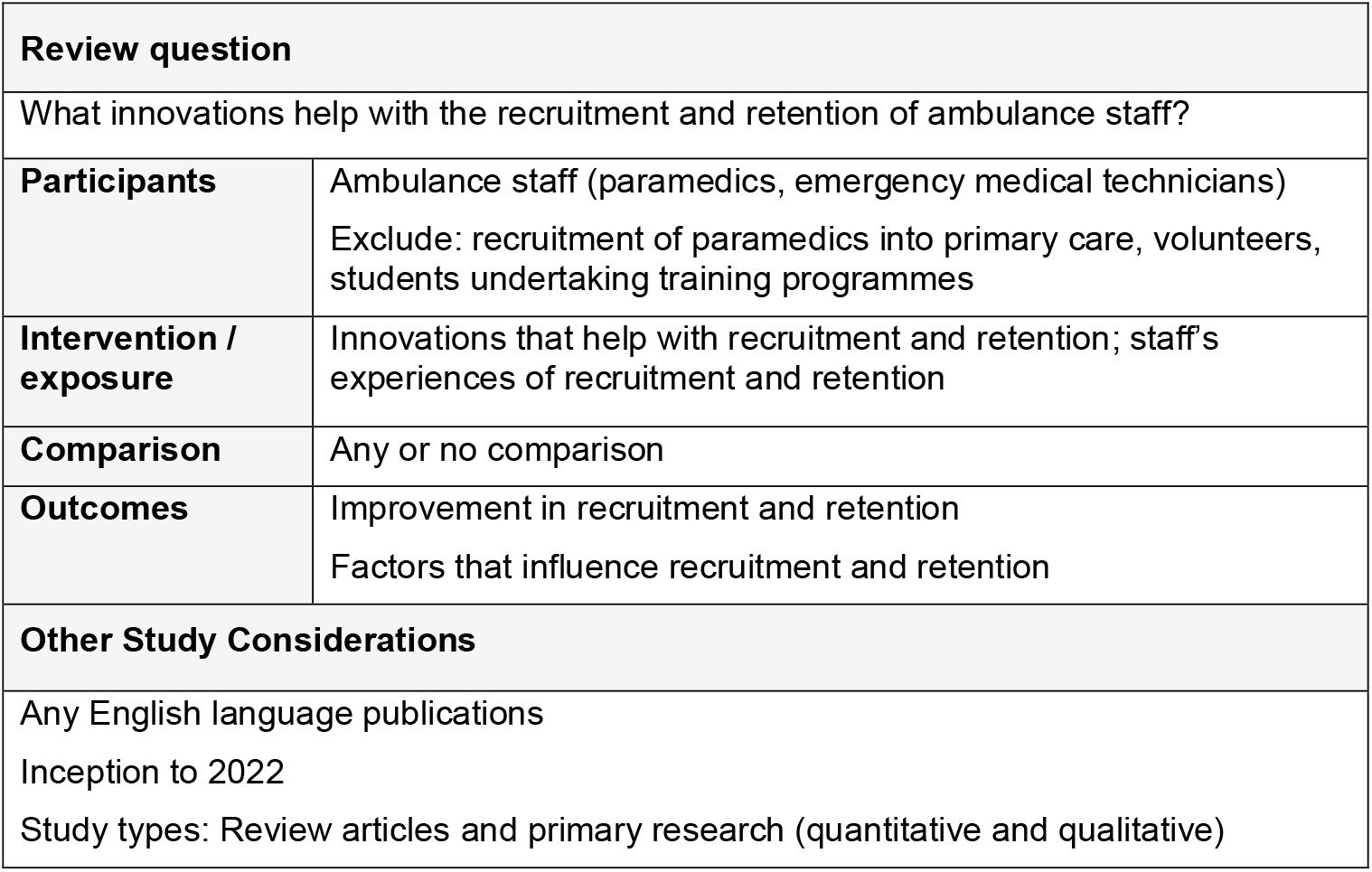

## 3. SUMMARY OF THE EVIDENCE BASE

### 3.1 Type and amount of evidence available

Six quantitative descriptive surveys and two qualitative studies were found that met the inclusion criteria. Six were USA studies, one UK and one Thai. A detailed summary of the included studies is presented in Section 6 (Table 2).

**Table 1.**
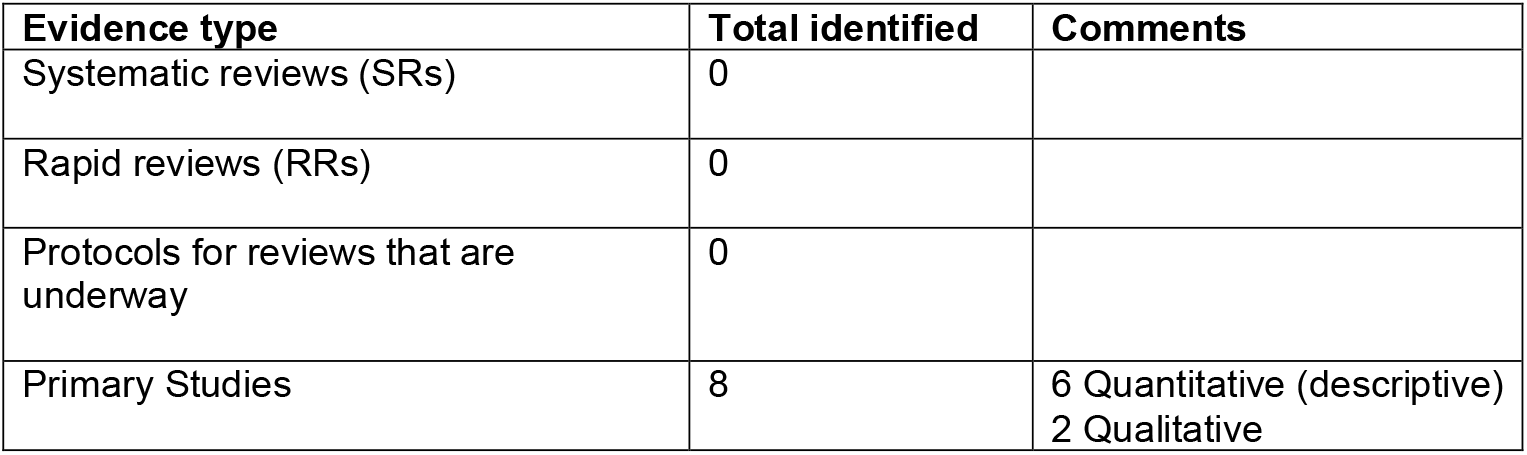
Summary of review evidence identified.

**Table 2:**
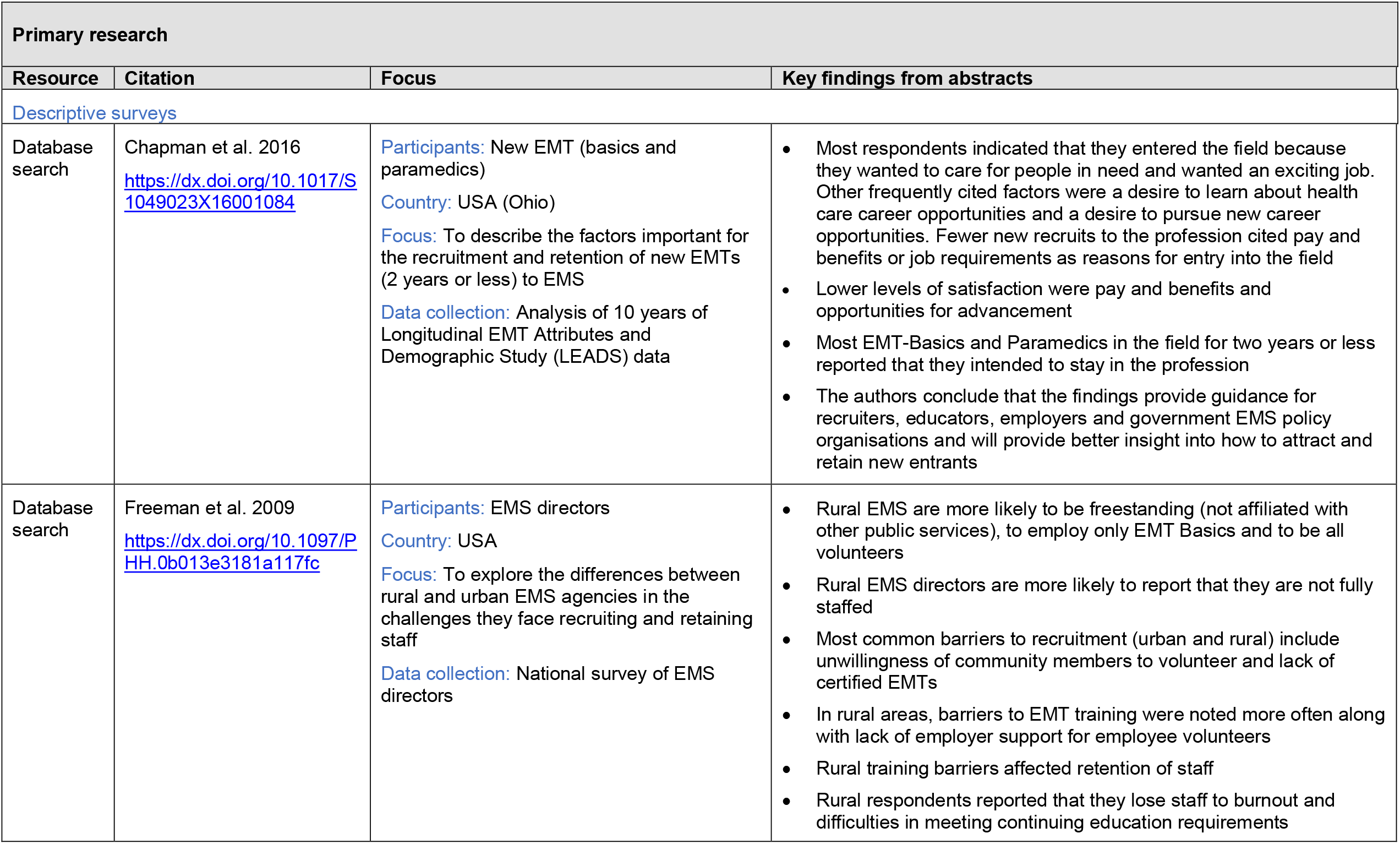
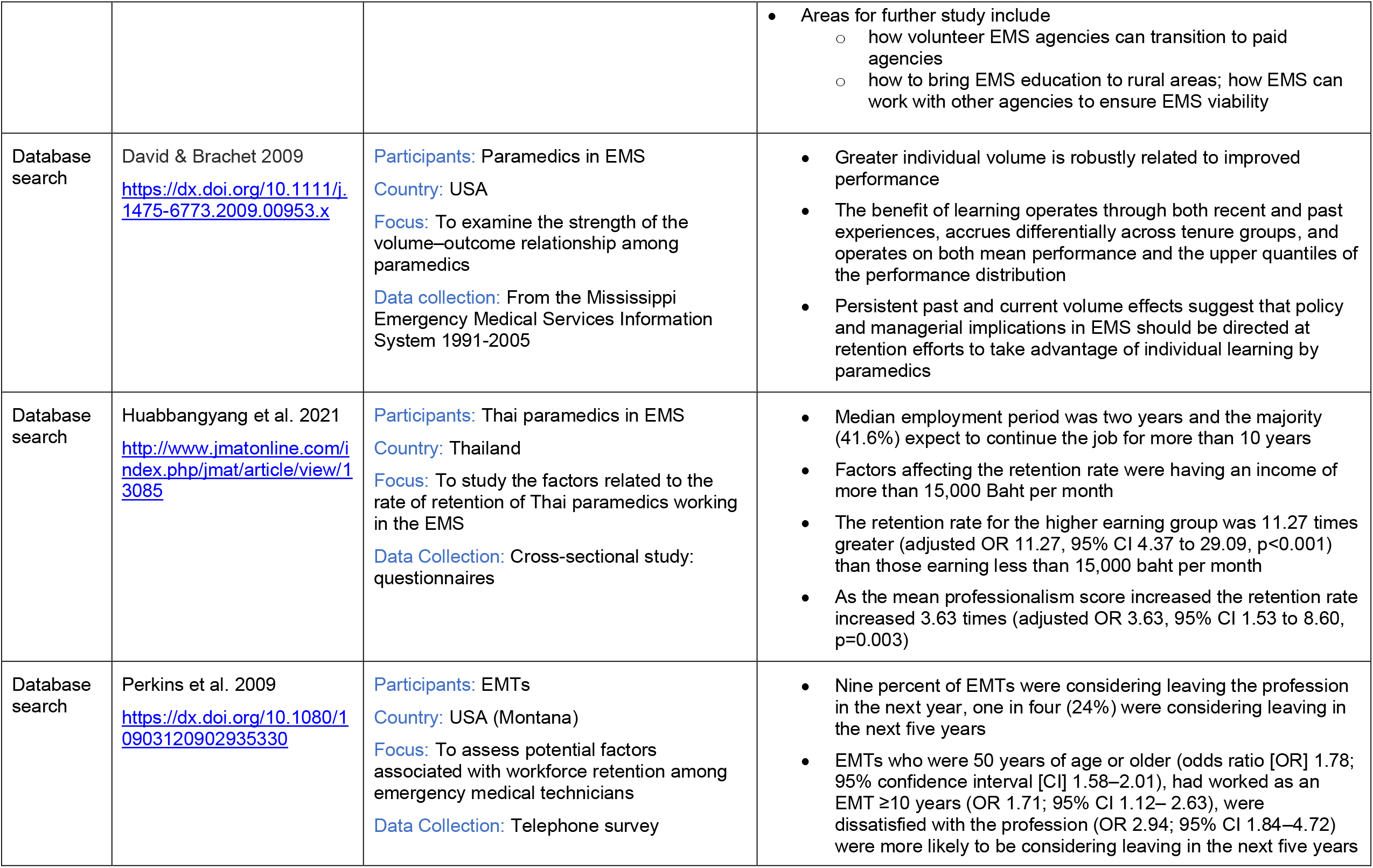
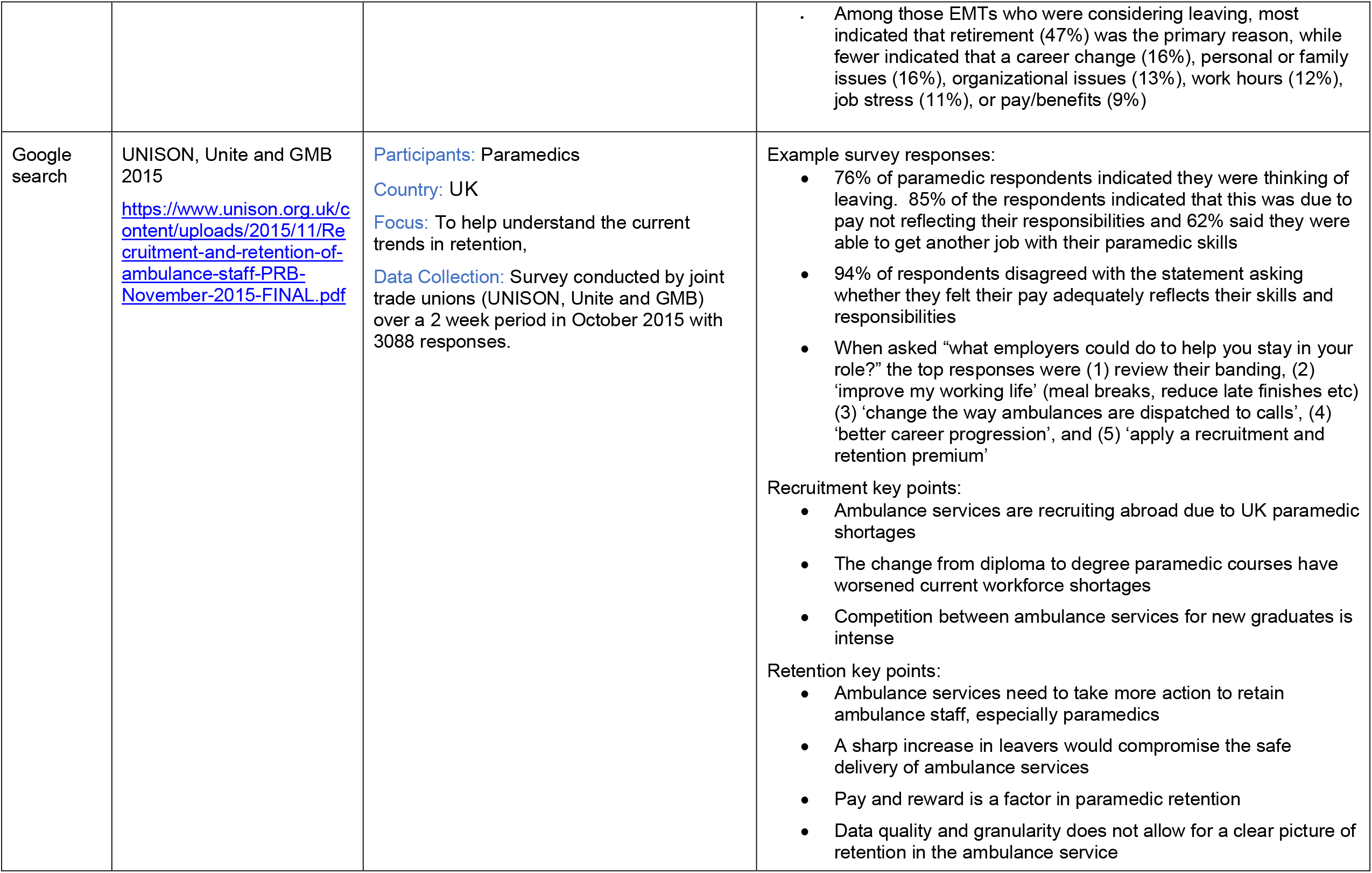
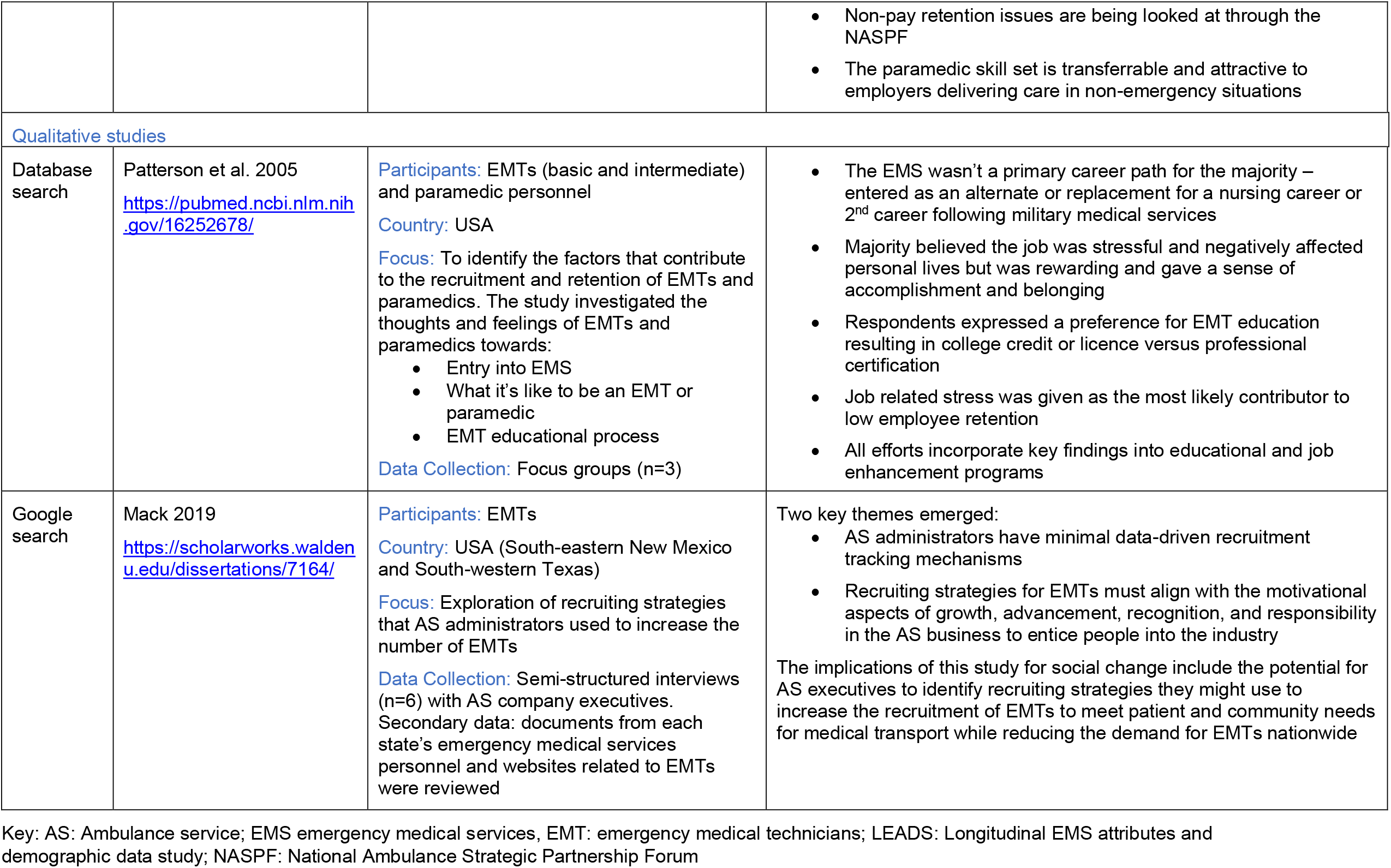
Summary of included evidence.

▪ A US study using linear and quantile methods to examine the strength of the volume-outcome relationship among paramedics where the authors hypothesised that by identifying the effects of individual learning on performance, they could also assess the value of paramedics’ retention (David & Brachet 2009).
▪ A US survey, using 10 years of longitudinal LEADS survey data, to describe factors important for the recruitment and retention of Emergency Medical Technician (EMT), basics and paramedics, new to Emergency Medical Services (EMS), that provided guidance for recruiters, educators, employers and governmental EMS policy organizations (Chapman et al. 2016).
▪ A National US Survey of local community-based EMS directors focused on the recruitment and retention of staff in both rural and urban EMS, reporting on staffing levels, affiliations, employee support and barriers to recruitment (Freeman et al. 2009).
▪ A cross-sectional study of paramedics working in the EMS in Thailand reported demographic details and expectations of the paramedic population as well as factors affecting the rate of retention (Huabbangyang et al. 2021).
▪ A US telephone survey of licenced EMTs assessed potential factors associated with workforce retention, focusing on whether respondents were considering leaving the profession, during what time period and why (Perkins et al. 2009).
▪ A UK NHS Pay Review was undertaken by the three main trade unions representing the ambulance service (UNISON, Unite and GMB 2015) and employers through the National Ambulance Strategic Partnership Forum (NASPF) to help understand the trends at the time in the recruitment and retention of ambulance staff including staffing levels, rates of attrition, pay, banding, workload, responsibilities, causes of stress and what employers can do to help (UNISON, Unite and GMB 2015).
▪ A qualitative doctoral study explored and reported on the recruitment strategies that AS administrators have used to increase the number of EMTs in South-East New Mexico and South-West Texas, USA (Mack 2019).
▪ A US qualitative study that identified the career paths of EMTs and paramedics and the factors contributing to their recruitment and retention (Patterson et al. 2005).

### 3.2 Key Findings

#### 3.2.1 Recruitment

##### From the US

▪ Paramedics **enter the field** because they want to **care for people in need** and **wanted an exciting job** (Chapman et al. 2016).
▪ There are **difficulties** reported in the **recruitment** of both volunteer and certified EMTs in **rural settings** (Freeman et al. 2009).
▪ The EMS **was not a primary career path** and was often seen as an alternate or replacement for a nursing career or second career following military medical services (Patterson et al. 2005).
▪ **Recruiting strategies** for EMTs must align with the **motivational aspects of growth, advancement, recognition, and responsibility** in the AS business to entice people into the industry (Mack 2019).

##### From the UK

▪ **Competition** between ambulance services for new graduates is **intense** with various incentives on offer including **favourable terms of appointment, golden hellos** and **relocation packages** (UNISON, Unite and GMB 2015).
▪ Responses from the NASPF show that recruitment campaigns are unsuccessful due to a lack of available paramedics in the UK. Most ambulance services are **running**
▪ **overseas recruitment exercises**, recruiting from **Europe and Australia** due to UK paramedic shortages (UNISON, Unite and GMB 2015).
▪ The change from **diploma to degree** paramedic courses have **worsened** current **workforce shortages** as new graduates are likely to come into the labour market with more debt from increased tuition fees making the private sector more attractive (UNISON, Unite and GMB 2015).

#### 3.2.2 Retention

##### From the US

▪ The main sources of **low job satisfaction** were **pay, benefits and opportunities for advancement** (Chapman et al. 2016).
▪ EMS managers’ report that they **lose staff** due to **burnout and difficulties in meeting continuing education requirements** (Freeman et al. 2009).
▪ In exploring the relationship of paramedic volume on performance it was found that **greater individual volume** improved performance particularly for paramedics with above median tenure (4 to 6.2 years). The authors suggest that current volume should be allocated across fewer paramedics as this can **improve retention** (David & Brachet 2009).
▪ **EMTs** who were **older**, those who had **longer tenure** in the profession, and those who were **dissatisfied** with the profession were more likely to be considering leaving the profession in the next five years (Perkins et al. 2009).
▪ The main reason for considering leaving within the next 5 years was **retirement. Other reasons** included a **career change and pay and benefits** (Perkins et al. 2009).
▪ **Job related stress** was given as the most likely contributor to low employee retention (Patterson et al. 2005).

##### From Thailand

▪ **Remuneration and professionalism** were the two factors related to the retention (Huabbangyang et al. 2021).

##### From the UK

▪ **Pay and reward** is a factor in paramedic retention (UNISON, Unite and GMB 2015).
▪ **Stress and workload** are a huge factor in staff leaving the ambulance service. (UNISON, Unite and GMB 2015).
▪ **Data quality** and **granularity** does not allow for **a clear picture** of retention in the ambulance service (UNISON, Unite and GMB 2015).
▪ **Non-pay** retention issues are being looked at through the **NASPF** (UNISON, Unite and GMB 2015)
▪ The paramedic **skill set** is **transferrable and attractive** to employers delivering care in
▪ **non-emergency situations** (UNISON, Unite and GMB 2015).
▪ When asked **“what employers could do to help you stay in your role?”** the top responses were:
  ▪ review their banding
  ▪ improve my working life (meal breaks, reduce late finishes etc)
  ▪ change the way ambulances are dispatched to calls
  ▪ better career progression
  ▪ apply a recruitment and retention premium

### 3.3 Areas of uncertainty

▪ There is a lack of evidence from the UK with only one survey published in 2015.
▪ There is limited international research available, particularly we could not find any studies that investigated the effectiveness of innovations that focused on improving recruitment and retention.
▪ Our previous work regarding recruitment and retention of NHS staff did not retrieve any research related to paramedics, indicating a clear gap in research (Edwards et al. 2022b, Edwards et al. 2022a).

### 3.4 Options for further work

▪ Conduct a more in-depth review that explores reasons for leaving the profession and/or factors affecting job satisfaction.
▪ Primary research in UK settings of paramedics’ and EMT’s recruitment and retention within the ambulance service is needed to provide up-to-date information about factors influencing recruitment and retention.
▪ Primary research could also focus on developing and evaluating strategies to help recruitment and retention of paramedics and EMTs.

## Data Availability

All data produced in the present study are available upon reasonable request to the authors

## Abbreviations

Acronym: **Full Description**
AS: Ambulance service
EMS: Emergency medical service
EMT: Emergency medical technician
HCPC: Health and Care Professions Council
LEADS: Longitudinal EMS attributes and demographic data study
NASPF: National Ambulance Strategic Partnership Forum
NHS: National Health Service
RES: Rapid evidence summary

## 5. RAPID EVIDENCE SUMMARY METHODS

General repositories of evidence reviews noted in our resource list were searched in October 2022. An audit trail of the search process is provided within the resource list (Appendix).

Searches were limited to English-language publications and included searches for primary studies as no secondary research relevant to the question was found. Search hits were screened for relevance by a single reviewer. Findings were tabulated for all relevant primary research identified in this process.

|  |  |
| --- | --- |
| **Date of Search** | October 2022 |
| **Search Concepts used** | Paramedic* OR ambulance* OR emergency medical OR emergency care assistant* AND recruit* OR retain* OR retention* |
| **Search Completed by** | Wales Centre For Evidenced Based Care |

## 6. EVIDENCE

A more detailed summary of included evidence can be found in Table 2.

## 7. ABOUT THE WALES COVID-19 EVIDENCE CENTRE (WCEC)

The WCEC integrates with worldwide efforts to synthesise and mobilise knowledge from research.

We operate with a core team as part of Health and Care Research Wales, are hosted in the [Wales Centre for Primary and Emergency Care Research (PRIME),](http://www.primecentre.wales/) and are led by [Professor Adrian Edwards of Cardiff University](https://www.cardiff.ac.uk/people/view/123022-edwards-adrian).

The core team of the centre works closely with collaborating partners in [Health Technology Wales](https://healthtechnology.wales/), Wales Centre for Evidence-Based Care, [Specialist Unit for Review Evidence centre](https://www.cardiff.ac.uk/specialist-unit-for-review-evidence), [SAIL Databank](https://healthandcareresearchwales.org/about-research-community/secure-anonymised-information-linkage-sail-databank), [Bangor Institute for Health & Medical Research/ Health and Care Economics Cymru,](https://www.bangor.ac.uk/medical-health-sciences/research) and the [Public Health Wales Observatory](https://phw.nhs.wales/services-and-teams/observatory/).

Together we aim to provide around 50 reviews per year, answering the priority questions for policy and practice in Wales as we meet the demands of the pandemic and its impacts.

### Director

Professor Adrian Edwards

### Website

https://healthandcareresearchwales.org/about-research-community/wales-covid-19-evidence-centre

## 8. APPENDIX: Resources searched during Rapid Evidence Summary

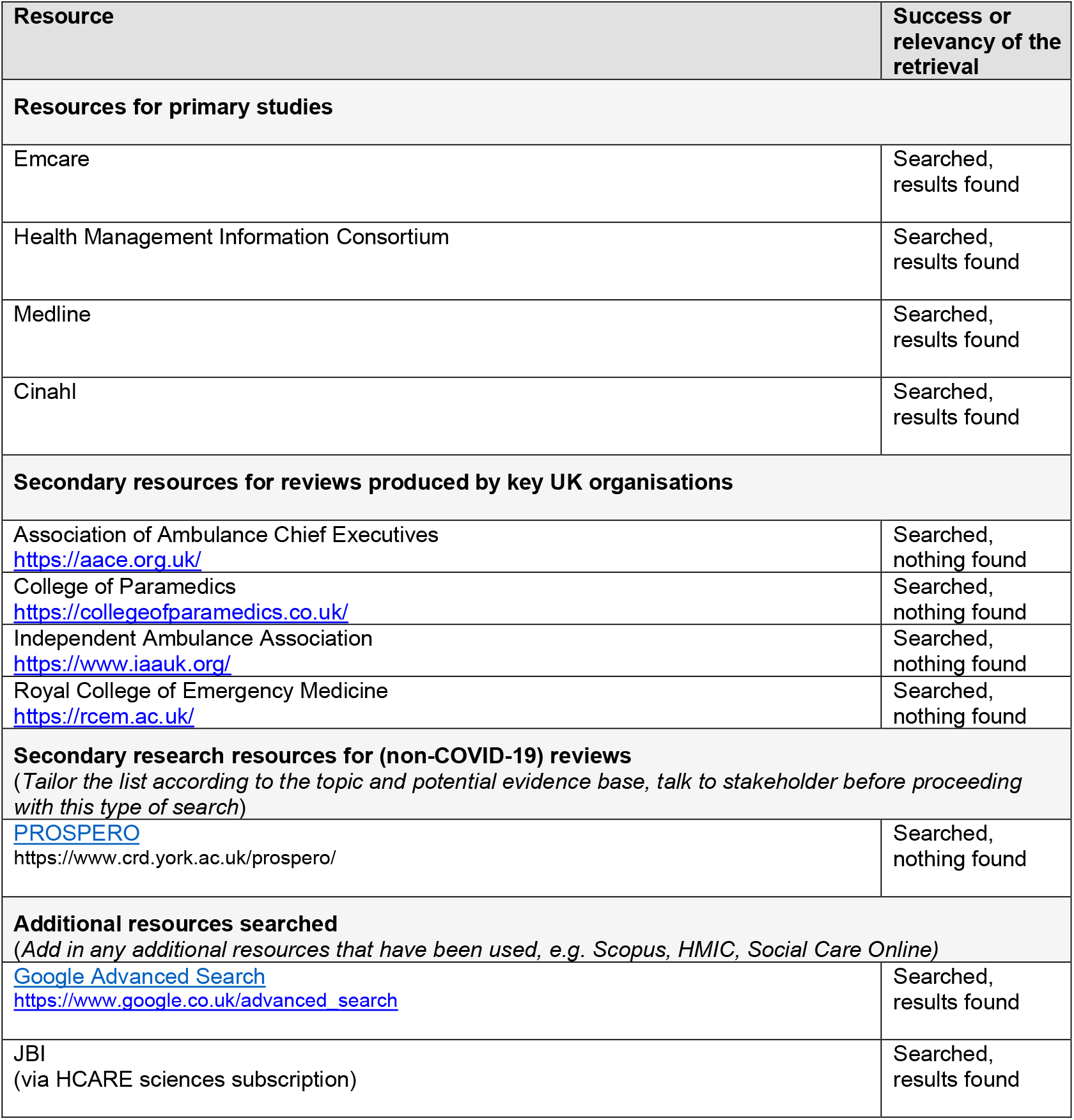

### Emcare

1. (paramedic* or ambulance* or emergency medical or emergency care assistant* or ems or emt).ti. (11555)
2. (recruit* or retain* or retention).ti. (25353)
3. 1 and 2 (40)
4. limit 3 to english language (36)

### Health Management Information Consortium (28/10/2022)

1. (paramedic* or ambulance* or emergency medical or emergency care assistant* or ems or emt).ti. (1186)
2. (recruit* or retain* or retention).ti. (1382)
3. 1 and 2 (7)
4. limit 3 to English language [Limit not valid; records were retained] (7)

### MEDLINE (28/10/2022)

1. (paramedic* or ambulance* or emergency medical or emergency care assistant* or ems or emt).ti. (18598)
2. (recruit* or retain* or retention).ti. (78472)
3. 1 and 2 (50)
4. limit 3 to english language (48)

### CINAHL

(TI (paramedic* or ambulance* or emergency medical or emergency care assistant* or ems or emt)) AND (TI (recruit* or retain* or retention)) (English language) (53)

| Database |  | Citations |
| --- | --- | --- |
| Medline |  | 48 |
| Emcare |  | 36 |
| HMIC |  | 7 |
| CINAHL |  | 53 |
|  | **Totals** | **144** |
|  | Duplicates | 68 |
|  | Title and abstract screening | 76 |
|  | Full text screening | 6 |

